# Variations in Non-Pharmaceutical Interventions by State Correlate with COVID-19 Disease Outcomes

**DOI:** 10.1101/2021.07.28.21261286

**Authors:** Annika J. Avery, Jiayi Wang, Xinyu Ma, Qingkai Pan, Elizabeth E. McGrady, Zongyuan Yuan, Yuqing Liang, Rebecca Nugent, Seema S. Lakdawala

## Abstract

The COVID-19 pandemic highlighted the lack of understanding around effective public health interventions to curtail the spread of an emerging respiratory virus. Here, we examined the public health approaches implemented by each state to limit the spread and burden of COVID-19. Our analysis revealed that stronger statewide interventions positively correlated with fewer COVID-19 deaths, but some neighboring states with distinct intervention strategies had similar SARS-CoV-2 case trajectories. Additionally, more than two weeks is needed to observe an impact on SARS-CoV-2 cases after an intervention is implemented. These data provide a critical framework to inform future interventions during emerging pandemics.

## Introduction

Starting in December of 2019, there were reports of a novel coronavirus (CoV) emerging in China; on March 11, 2020, the World Health Organization (WHO) declared COVID-19 a pandemic. In response, many countries around the world began implementing non-pharmaceutical interventions (NPI) and providing guidance to keep their citizens safe, with several countries (such as China, South Korea, Singapore, Japan and in Europe) doing so in late February and March of 2020 (*1-3*). In contrast, the United States federal government did not implement and oversee any federal NPIs to restrict transmission of COVID-19 during all of 2020. Instead, individual states carried out the responsibility of implementing restrictions and creating reopening plans with their respective state-level resources (*4*).

This necessary action from US state and local governments resulted in a variegated approach to managing an emerging pandemic and over 600,000 deaths in the US as of July 2021 (https://coronavirus.jhu.edu/). In the face of this unfathomable loss, there exists an opportunity to understand how specific NPIs may affect transmission of COVID-19 in the United States. We aimed to characterize the effectiveness of NPIs implemented at the state level at reducing COVID-19 transmission as well as compare the overall effectiveness of NPI implementation strategies across states.

To explore the dynamics of NPI implementation and changes in daily COVID-19 cases, we created an interactive dashboard (www.phightcovid.org) that displays state-specific time series with normalized daily COVID-19 cases to compare the potential impact of NPIs across states. This dashboard is periodically updated to incorporate more recent case counts and NPIs. Clustering the estimated basis-spline models underlying the state time series revealed geographical groupings, indicating that similar SARS-CoV-2 case trajectories may be based on state location in addition to NPIs. Finally, we observed a statistically significant negative correlation between COVID-19 deaths and state NPI score, solidifying the importance of implementation of public health interventions.

### Heterogeneity in State NPI restrictions

To capture the heterogeneity in protective measures implemented by each state, including the District of Columbia (DC), we collected state mandated COVID-19 NPI orders from each governor website for one year (March 11, 2020-March 31, 2021). NPI orders for the following five categories were included in this study: stay-at-home orders, non-essential business restrictions, indoor gathering limitations, restaurant/bar restrictions, and mask/face covering mandates. Daily cumulative NPI scores were calculated for each state based on the stringency of each intervention in these five categories (see methods for score rubric). Fig. 1 highlights the heterogeneity in the NPIs implemented by each state, at three distinct times during the pandemic. A time-lapse movie of the NPI map with daily COVID-19 cases is provided in movie S1. Interestingly, implementation strategies varied by state with some states directly placing orders on all residents, while others delegated the responsibility for ordering NPIs to county or city governments (fig. S1A). In some cases, states initially placed statewide orders on all residents, but as the pandemic progressed, implementation of interventions was moved to a county or city level. Fig. S1 provides a snapshot of states that implemented statewide, county-level, or city-level NPIs, note that the various levels of NPI implementation were not exclusive. In some states, statewide NPIs were implemented for all residents, but individual counties would extend or modify the NPIs (fig. S1). To explore the accessibility of NPI guidelines per state, we additionally measured the time it took for a new user to find NPI information for each state (fig. S1B). We observed that information for the majority of states could be found in under five minutes (fig. S1B).

**Figure 1:**
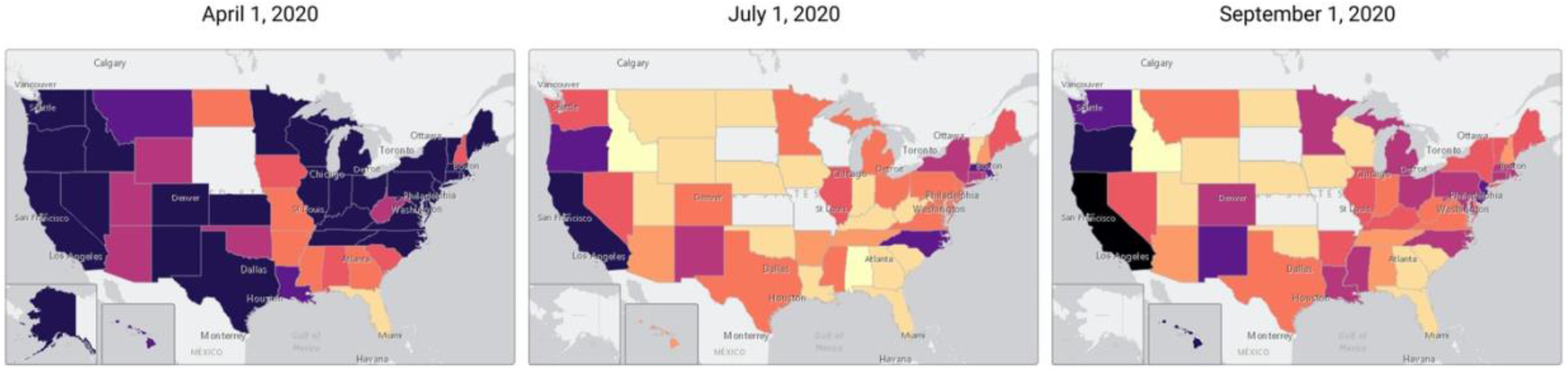
Spatial temporal maps showing state NPI score over time display heterogeneity in state responses. Each restriction was out of a maximum of one point. States had the maximum score of five points when all five categories had restrictions at 100%, (i.e., either full closure or extreme limitations, depending on the category). The darker the color of a state the stronger the NPI restrictions (and the higher the score).

All states, except for South Dakota where there weren’t any mandatory statewide NPIs, implemented a different combination of interventions statewide during the pandemic (www.phightcovid.org). Initially, 43 states implemented stay-at-home orders, of which 93% were lifted by June 15, 2020. Restaurant/bar closures were implemented in 49 states (including DC) between March 15 and April 3, 2020. These states, except Missouri, eased restaurant/bar restrictions for the first time between April 24 and June 15, 2020, with the largest proportion doing so between May 9 and May 19, 2020. Missouri did not ease this NPI, rather the closure was completely lifted on June 16, 2020. Between June 1, 2020, and January 2, 2021, 41 states and DC reissued restaurant/bar restrictions for some duration of time before again easing them. A similar scenario of alternating between easing and re-issuing mandates occurred with indoor gathering limitations and non-essential business restrictions. Temporal restrictions by state are available in an interactive format on www.phightcovid.org/graphs.

### SARS-CoV-2 case trajectories cluster by geographical region

To compare the implementation of various NPIs per state over the last year to the trajectory of COVID-19 cases, we created time series graphs, (including a seven-day rolling average) for each state as displayed in Fig. 2 and fig. S2. In Fig. 2, Maryland and Tennessee are highlighted as representative states that implemented strong interventions throughout the past year (Maryland) versus states that had implemented fewer interventions (Tennessee) (table S1). The implementation dates of the statewide government NPIs are indicated on the graphs. As NPIs were implemented and eased, the NPI score changed based on the heuristic NPI scoring rubric, indicated by the colored horizontal bar (table S2). While most states, including Maryland and Tennessee, implemented NPIs early in the pandemic, Tennessee was far less stringent with NPIs in the fall of 2020 when cases began to rise. At the beginning of October, Tennessee lifted all NPIs (NPI score = 0), and in December the state experienced record new daily SARS-CoV-2 cases during the fall/winter wave. Tennessee briefly reissued indoor gathering limitations of ten people from December 21, 2020 to January 19, 2021 to curtail the spread of COVID-19 (table S1). Many states, like Maryland, eased restrictions during the summer, but reissued multiple restrictions in the fall as cases began to increase. For example, Maryland reissued restrictions in November that limited indoor gatherings to 25 people as well as restrictions on restaurant/bar occupancy, alcohol curfews, and occupancy limits at non-essential business (table S1). Like most states, Maryland and Tennessee experienced record new daily COVID-19 cases in winter months (December 2020 to January 2021), although the amplitude of the peaks differed. The COVID-19 peak of new daily COVID-19 cases was 271% greater in Tennessee compared to Maryland. Across all 50 states there was variation in SARS-CoV-2 case trajectories, but states with consistent NPI restrictions throughout the year had smaller fall/winter peaks (www.phightcovid.org/graphs). This observation would suggest states may have similar epidemic curves related to the strength of the NPI restrictions implemented in each state.

**Fig 2:**
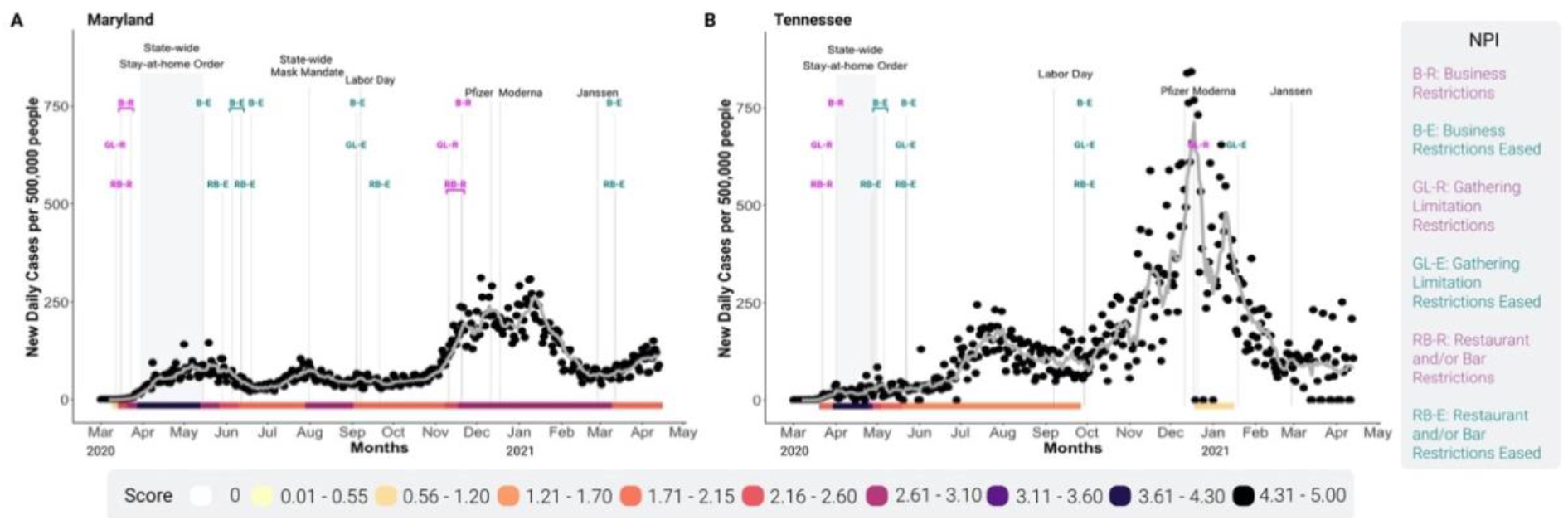
SARS-CoV-2 cases and non-pharmaceutical interventions are distinct between states. Time series graphs for Maryland (A) and Tennessee (B) from March 11, 2020, through March 31, 2021. Daily SARS-CoV-2 cases were normalized to state population represented per 500,0000 people (black circles), and the seven-day rolling average is indicated by the grey line. State NPI score per day is indicated by the colored bar along the x-axis. The NPIs are labeled with the category abbreviation, and colored as magenta if restricting, or teal if easing. The duration of the stay-at-home order is indicated by the grey band; black vertical lines represent implementation of statewide mask mandate, labor-day, and each vaccine FDA approval dates.

To identify similarities between state SARS-CoV-2 case curves, we estimated a basis spline (B-spline) model for every state (*5*). Each estimated B-spline is a weighted piecewise combination of 15 polynomials, connected at “knots”. Examples of the estimated B-splines and their SARS-CoV-2 case curves for three states (Texas, Georgia, and Maine) are included in fig. S3A-F; these panels show how the different polynomials are combined into the overall estimated spline. Estimated splines and weight coefficients are closely related in states with similar underlying case curves. We compared the estimated state splines by using the K-means algorithm to cluster similar sets of weight coefficients, identifying groups of states with similar SARS-CoV-2 case trajectories. Returning to our three example states, we see that the coefficients for two states in the same cluster (Texas, Georgia) are more similar than those in a different cluster (Maine) (fig. S3D&E compared to F).

Standard best-fit criteria for K-means and repeated simulation identified a set of seven clusters of states, each with a distinct case curve shape shared across each group. The most common clustering is depicted in (Fig. 3A). Interestingly, the clusters demonstrate that states grouped geographically even though no geographic or NPI information was included when identifying the clusters, indicating that neighboring states tended to have similar case curve trajectories even if their NPI interventions differ (Fig. 3A).

**Fig 3:**
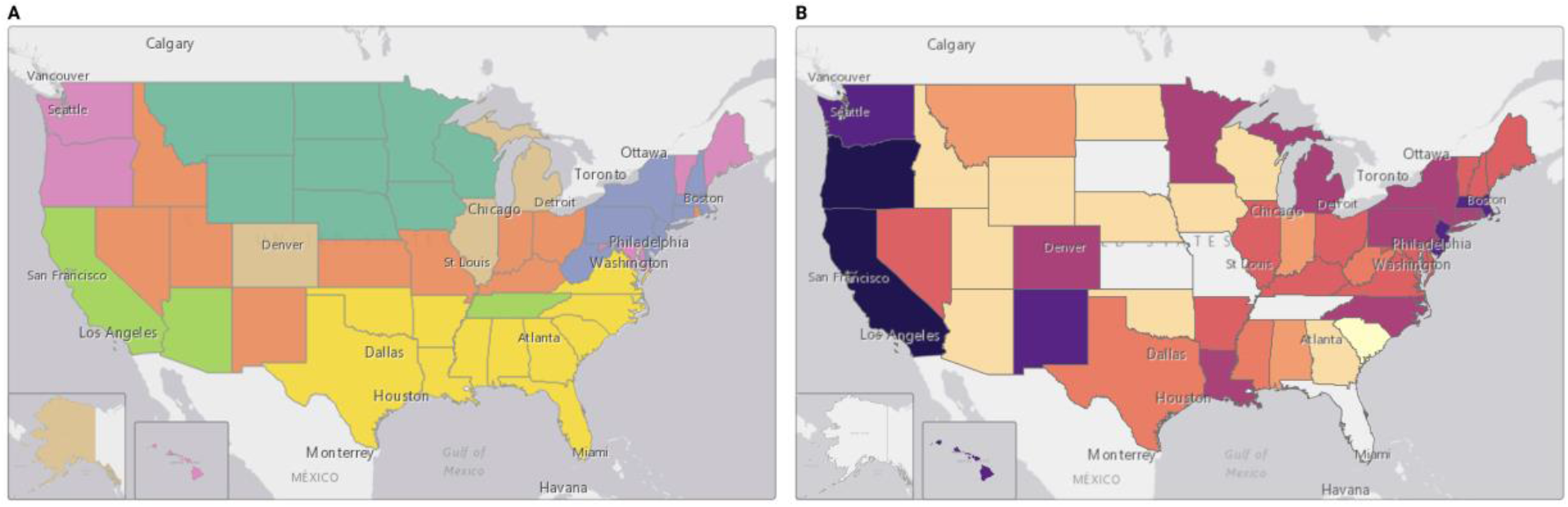
Grouping of states geographically based on SARS-CoV-2 case trajectories. (A) Map showing the most common state groupings produced in our B-spline model clustered at 7, with 15 knots for 20 simulations. (B) Map showing the median state NPI score for each state. States were colored according to the colored bar shown in Fig. 2.

An assessment of state median NPI score from March 2020 to March 2021 is depicted in Fig. 3B; surprisingly, a similar clustering pattern was not observed between these two maps. For example, Minnesota and Wisconsin have distinct state median NPI scores but fall within the same K-means clusters (Fig. 3A and B), suggesting that implementation of various NPIs may not solely dictate the trajectory of SARS-CoV-2 cases. Aside from geographical location, environmental conditions such as humidity and temperature are thought to contribute to the seasonality of influenza viruses (*6*). Recently, humidity and temperature have been implicated in SARS-CoV-2 spread (*7*). However, the identified clusters may not correlate directly to environmental conditions, rather it could be driven by interstate travel patterns, similarities in state demographics, or when specific interventions were implemented(*8, 9*). Mobility and social behavior have also been linked to transmission of COVID-19 and may also influence the spread between states(*10-12*). These potential confounders all require further investigation.

### Impact of NPI on case change occurs at greater than 2 weeks

Effective implementation of NPI requires an understanding of the impact of NPIs on the trajectory of SARS-CoV-2 transmission. During the COVID-19 pandemic, discussion around the impact of interventions focused on case changes less than two weeks after an intervention. This included both easing or a restricting of an NPI (*13, 14*). However, that may not be a realistic time lag between NPI and case change. To examine this relationship, we graphed the number of cases at the time of an intervention versus the number of cases some point in the future, either 2-, 4-, and 5-weeks after an intervention (Fig. 4). Michigan was chosen as a representative of states that implemented many NPI over the course of the last year. By examining the NPIs within Michigan, we found that cases 2-weeks after either an easing or restricting NPI were similar to cases when this NPI was implemented, as the spots are still along the line of identity (x=y). The farther away from the line of identity the greater the case change; we anticipate that as cases raise after lifting of restrictions the cases will increase and move toward the red circle. In contrast as cases reduce after the implementation of an NPI, cases will decrease and move toward the blue circle. Fig. 4B and 4C examine case change after NPIs at 4-and 5-weeks later. Cases 4 weeks after a NPI is eased have increased such that the red easing restrictions are in the red circle (Fig. 4B). In contrast, cases 4 weeks after a restrictive NPI is implemented have lower cases, but they continue to decrease 5 weeks after the restrictive intervention (Fig. 4C). The observations made in Michigan pose interesting implications regarding the duration of time an NPI should be left in place until it becomes effective or when NPIs should be implemented. This analysis did not distinguish between the type of NPI and impact on cases, which could impact the effect on cases and requires further exploration.

**Figure 4:**
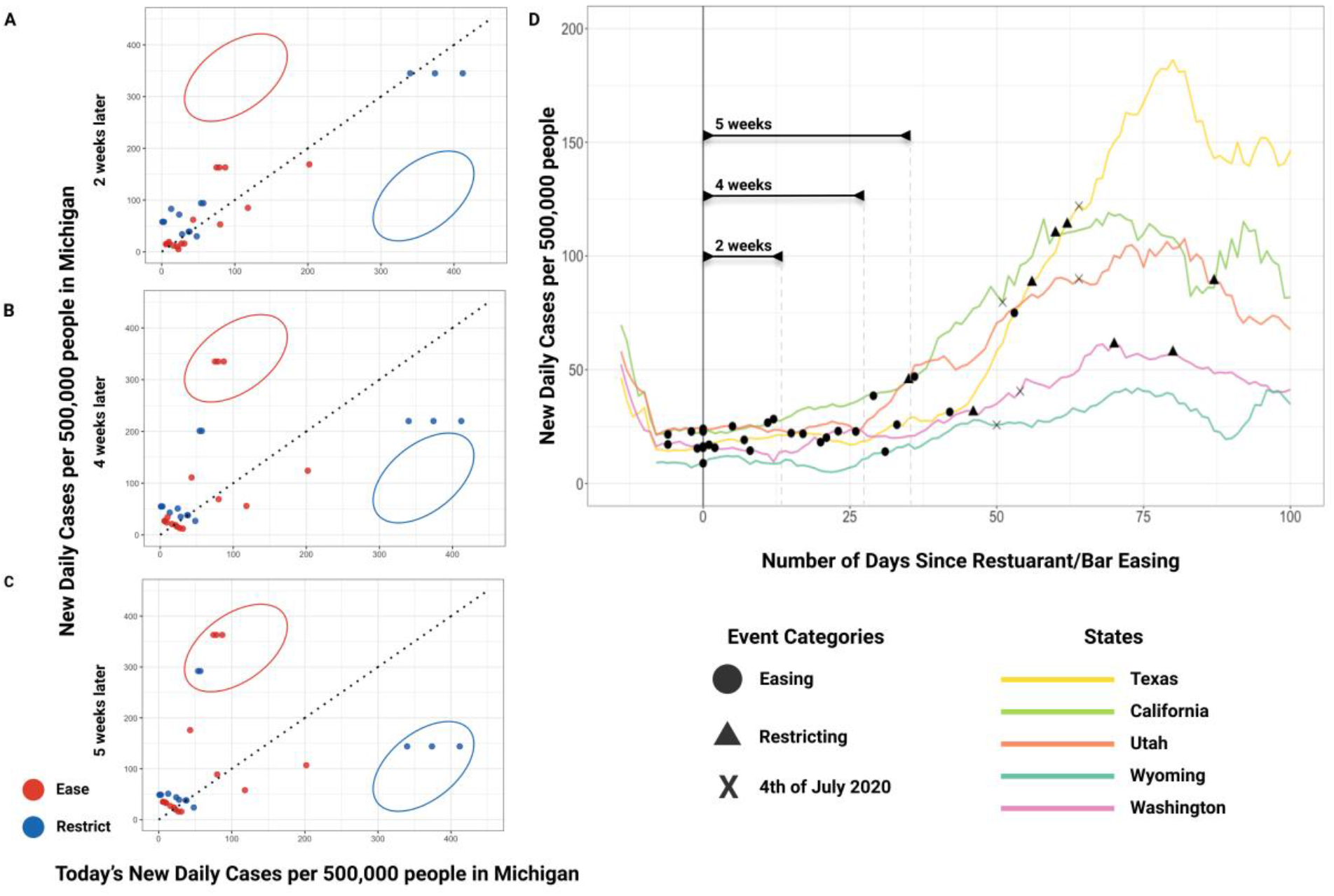
More than two-weeks is needed before a NPI has an impact on case change. Case change between the date a restrictive NPI intervention was issued two (A), four (B) and five (C) weeks later for Michigan. Interventions are represented by the points, with easing shown in red and restricting in blue. The circled areas show where the points will be located if there is a considerably greater (red), or less (blue) number of cases at 2, 4 and 5 weeks later. The line of identity, where x=y is the diagonal dashed line in the center of the graph. (D) Overlapping time series graphs showing the number of days since the first time a restaurant/bar easing was issued after May 2020, represented by day 0 on the x-axis. NPIs that were eased are represented by circles. NPIs that were restricted are represented by triangles. The Fourth of July is marked with an X for reference.

To assess the impact of a specific type of NPI, we overlayed the SARS-CoV-2 case trajectories from multiple states on the day a specific intervention was implemented. Using states from multiple different clusters as shown earlier (Fig. 3A), we graphed the lag in days from the first-time restaurant/bar restrictions were eased after May 2020. Two weeks after the first restaurant and bar easing in May all states were relatively similar in that all experienced little case change. However, between two and four weeks, and even more so between four and five weeks, cases increased. This analysis further supports the observation that a lag of more than 2-weeks is needed before a change in cases after an NPI is eased or implemented (Fig. 4D).

### COVID-19 disease outcome correlates with NPI score

To determine whether the implementation of various NPIs and their stringency impacted COVID-19 outcomes in each state we compared the median state NPI score for each state to SARS-CoV-2 cases and COVID-19 deaths. A linear regression of cumulative SARS-CoV-2 cases normalized to state population and state median NPI score revealed a statistically significant negative relationship (Fig. 5A, p <0.0001). This suggested that greater NPI restrictions led to lower cases, similar to what was reported by other recent modeling approaches (*15, 16*). Since case reporting has been found to vary by state (*17, 18*), we compared state median NPI score to cumulative COVID-19 deaths, normalized to state population, from June 2020 -March 31, 2021 to remove the potential confounder of variable case reporting (Fig. 5B). Linear regression indicated that median state scores were negatively correlated with cumulative COVID-19 deaths (p=0.02), indicating that states with more interventions had reduced mortality rates in addition to fewer cases.

**Figure 5:**
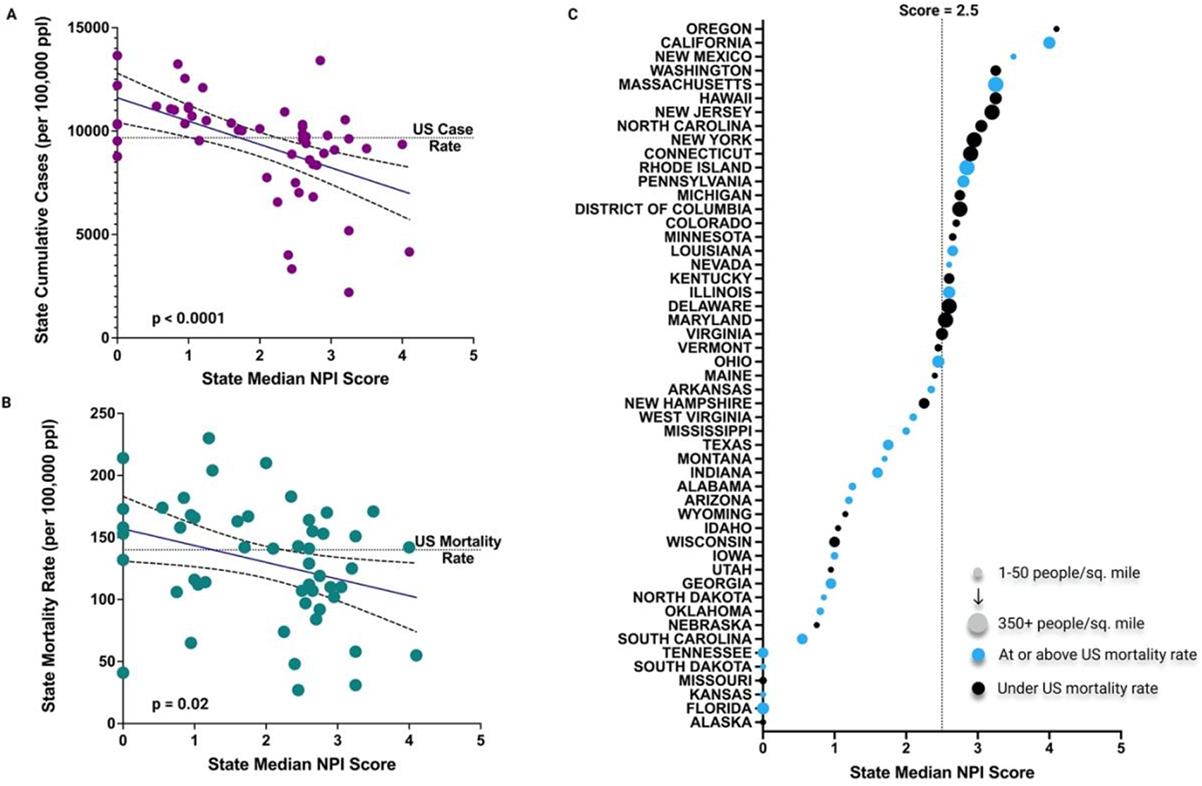
Higher state NPI scores are associated with lower COVID-19 cases and deaths. (A) Linear regression of state median score and SARS-CoV-2 case rate March 2020-April 2021. State cumulative cases are normalized to 100,000 by state population. Dashed lines show the 95% confidence interval. (B) Linear regression of state median score and SARS-CoV-2 mortality rate June 1, 2020-April 13, 2021. State mortality rate is normalized to 100,000 by state population. Dashed lines show the 95% confidence interval. (C) Scatter plot of all state median scores, US mortality rate and population density. Dashed line along the y-axis highlights median score of 2.5. States are colored as blue if their COVID-19 mortality rate was at or above the US COVID-19 mortality rate. State population density is the number of people per square mile and is represented by the circle size. States with a large circle size have a high population density. We grouped population density as follows, extra-small: 1-50 people/sq. mile; small: 50-100 people/sq. mile; medium: 100-200 people/sq. mile; large: 200-350 people/sq. mile; extra-large: 350+ people/sq. mile.

While the trendlines are clearly significant between NPI score and SARS-CoV-2 cases or COVID-19 deaths, there are a number of states that reside outside of the 95% confidence interval (Fig. 5A and B). In particular, we were surprised by the states that had high NPI scores, but a similar case or cumulative death value as states with much lower NPI scores, suggesting that there may be an optimal NPI restriction threshold. To define the ‘goldilocks’ or ‘just right’, of NPI combinations, we compared the states with cumulative cases below the national cumulative case or death average for common interventions, including mask orders, some level of restaurant/bar restrictions, and gathering limitations. Using our rubric, states with all three intervention combinations at some stringency level for a prolonged period of time during the pandemic would have a median state NPI score of ∼2.0-3.0, and a greater likelihood of lower mortality and fewer cases than states without these interventions. For example, Maryland, shown in Fig. 2A, had a median score of 2.55 and had the 12^th^ lowest mortality rate out of all states.

Overall, 45% of states had a median NPI score above 2.5 and of these only 8 out of 23 had a mortality rate above the national average. In contrast, in states that had a median score below 2.5, 18 out of 25 had a mortality rate higher than the US average mortality rate (Fig. 5C). It is interesting to note that in states with the highest population density, 14 of 16, had a median score above 2.5, indicating stronger restrictions. In contrast, only 5 out of 24 states with the smallest population densities had median NPI scores above 2.5. To compare how population density may contribute to COVID-19 mortality in the context of NPI orders, a regression between average cumulative deaths per state by population density was performed for states with median NPI. Surprisingly, we did not find a statistically significant relationship between population density and cumulative mortality rate from June 2020 - March 31, 2021, for state median NPI scores either above or below a median score of 2.5 (fig. S4).

## Discussion

During the course of the COVID-19 pandemic a number of studies have attempted to examine the effectiveness of NPIs on SARS-CoV-2 cases around the world and in the US. These studies focused on NPIs implemented early in the pandemic (prior to June 2020) and found that NPIs reduced SARS-CoV-2 cases and mobility with both real and modeled data (*1, 2, 10, 15, 18-21*). Comparison of 190 countries from January 23, 2020 to April 13, 2020 found that countries with NPIs (specifically: face mask, quarantines, social distancing, and travel restrictions) may have had a lower R_t_ value, suggesting a decrease in secondary SARS-CoV-2 transmission events in these spaces (*1*). However, this broad view does not integrate social behavioral variations or climate impacts that could also influence SARS-CoV-2 transmission. In other studies focused on U.S. counties or states, it was found that NPIs reduced mobility of the population and increased the doubling time of SARS-CoV-2 infections (*18, 19*). These studies again examined NPIs implemented early in the pandemic and could not account for issues with SARS-CoV-2 testing or the multiple rounds of issuing and easing interventions throughout the course of the past year. In contrast, our study includes data from all 51 U.S. states (including DC) and encompasses the NPIs implemented from March 2020 to April of 2021, to incorporate multiple waves of SARS-CoV-2 infections in states.

Taken together our results indicate that state implemented NPIs can lead to less COVID-19 cases and mortality. While it was previously thought that more NPIs led to a better outcome of disease burden (*15*), we found that even a moderate amount of restrictions can have a substantial impact on lowering COVID-19 transmission. In addition, we observed a clear geographical impact on SARS-CoV-2 trajectories, and it is likely that consideration of geographical neighbors should be considered when designing future pandemic NPI plans. Finally, additional analysis into the impact of interventions in each state is needed to account for the numerous confounders that limit this type of analysis. A more refined analysis examining the impact of interventions, when they were implemented, the state of the disease at time of implementation, the strength of interventions, and implementation of similar interventions in neighboring states, is needed.

## Methods

### Data Collection

COVID-19 case and death data were obtained from the COVID-19 Data Repository by the Center for Systems Science and Engineering (CSSE) at Johns Hopkins University. For JHU CSSE COVID-19 Data see https://github.com/CSSEGISandData/COVID-19. Using this data, we calculated the new daily cases/500,000 people for every state by normalizing a state’s new confirmed cases to the state population then multiplying by 500,000.

NPI intervention data was collected from a variety of sources and evolved over time. Data was collected manually from state government and governor websites. Data obtained on these websites was found in press releases, FAQ pages, reading through executive orders, or in the most accessible format, through an informative interactive portal detailing current restrictions and any changes from past restrictions. Data obtained through various news sources or governor’s twitter page, was validated through a search for an official state government or governor announcement or order.

Implementation of NPIs varied between statewide or county specific order, which will impact the ability for residents to obtain this information and their awareness of the NPI in their region. To address the accessibility of the NPI orders in place for each state, we measured the time it took to 1) find state issued COVID-19 restrictions and 2) determine if state government orders apply statewide or county-specific. Additionally, we identified if there were county government issued NPIs and the specific stringency of the orders. The search process was consistent for all 50 states and took place Feb 17, 2021. Starting at google.com the keywords “[state of interest] covid restrictions” was queried. The most relevant (state government page or governor page) first result was chosen. After entering the website, a process of looking for the above-mentioned questions was carried out. The time was recorded upon finding each of the questions of interest. Furthermore, if the site was exceptionally easy to navigate and the current restrictions in place were found in less than a minute without downloading and opening additional files, the state was noted as being outstanding in user accessibility (n=16, CA, CO, CT, DE, DC, ID, IL, MI, MN, NJ, NM, OH, OR, PA, RI, and WA). Although states without any NPI orders or no detail on the NPI orders state were also easy to navigate. In situations such as this the state was not marked as being outstanding in user accessibility due to their lack of information displayed at all (AR, FL, and SD). Details of the time, sources, and notes per state for this analysis is available as table S3.

### Scoring System

In order to compare the stringency of interventions implemented in each state, we developed a detailed scoring system. The system scores on the following five categories: Stay at Home Orders, Non-Essential Business Restrictions, Indoor Gathering Bans, Restaurant/Bar Restrictions and Mask/Face Covering Mandates. The highest score possible for each of the five categories is 1.00, with the collective highest score possible being 5.00. The scoring criteria are presented in table S2 and briefly described here:

- Stay at home orders had a score impact of 1.0 when issued and -1.0 when removed.
- Non-essential business sectors that we impacted the score on are as follows: retail (+/- 0.25), hair salons/barbershops (+/- 0.10), personal care services (+/- 0.10), gyms/fitness centers (+/- 0.2), indoor entertainment (+/- 0.2). Each of these business sectors could have between 0 and their max score depending on the level of the restrictions in place. For example, if hair salons were open but only at 50% occupancy, the score impact would be 0.05).
- Indoor gathering bans primarily applied to indoor private social gatherings, however, as states eased restrictions, indoor gathering sizes described were more closely associated with larger private events. See table S2, category 3 for the scores associated with gathering sizes.
- Restaurants and bars being completely closed resulted in a score impact of 1.0. The breakdown of the 1.0 is as follows: restaurant outdoor dining (+/- 0.1), restaurant indoor dining (+/- 0.45), bar outdoor service (+/- 0.1), bar indoor service (+/- 0.25), restaurant and/or bar having any distancing or sanitation practices (+/- 0.05) and restaurant and/or bar having a service or alcohol curfew (+/- 0.05). See table S2, category 4, for further details.
- Mask and facial covering restrictions had a score impact of 1.0 if they were required in both indoor and outdoor spaces and a score impact of 0.8 if they were only required indoors.

Scoring of states that adopted a state government issued county risk-level metric (CA, CO, HI, IL, IN, NM, ND, OR, UT, WA) was more complicated than just scoring for the state as by itself. In these states the NPIs scored on were whatever they were for the majority risk level out of all counties. For example, if Oregon had 16 counties at low, 6 at moderate and 2 at high risk levels, then the entire state would be considered as being at a low risk level. The low risk level NPIs would be recorded and the state as a whole scored scoring accordingly.

### Time Series

Time series graphs were generated for each state to visualize an individual state’s precise interventions (either restrictive or easing), NPI score (calculated based on the system described above), new daily SARS-CoV-2 cases, and seven day rolling average line. Daily SARS-CoV-2 cases were normalized to state population (based on the U.S. census bureau 2017 population estimates) and presented per 500,000 people. Interactive time series graphs are displayed on www.phightcovid.org, where the user can hover over each intervention and see the description of the type of intervention. The code to generate each of the time series is available on the phightcovid Github repository (see code availability section).

### Lags

Lag graphs were created for gaining a better initial understanding of the time needed between when an NPI is implemented and when cases begin to change as a result. Using the programming language R we plotted today’s new daily cases per 500,000 people on the x-axis and the new daily cases per 500,000 people at a lag of 2-, 4, and 5-weeks later, on the y-axis. We then use larger circles to highlight where it was expected to see the points if there was a considerable case change. R code for the lag graphs can be found in our Github repository

### Spline model and clustering

Basis spline models were generated with 15 knots per state. K-mean clustering of the 15 B-spline coefficients (adapted from Rory Michelen, https://github.com/RoryMichelen/Medium-Articles/blob/master/) was used to compare the SARS-CoV-2 case trajectories between states. Code for generation of splines and clustering can be found at the Github repository.

## Data Availability

The data analyzed in this manuscript, including the NPI scores per state, all code developed to generate time series graphs, lag graphs, and spline models as well as a collection of many pdfs and screenshots of the raw data sources organized by state are available on Github at https://github.com/Lakdawala-Lab/PHIGHTCOVID_StNPI_Publ2021. Case data for time series plots was obtained every other month from March 2020 to April of 2021 from the COVID-19 Data Repository by the Center for Systems Science and Engineering (CSSE) at Johns Hopkins University, available on Github at https://github.com/CSSEGISandData/COVID-19 (17).

## Code and data accessibility

The data analyzed in this manuscript, including the NPI scores per state, all code developed to generate time series graphs, lag graphs, and spline models as well as a collection of many pdfs and screenshots of the raw data sources organized by state are available on Github at https://github.com/Lakdawala-Lab/PHIGHTCOVID_StNPI_Publ2021.

Case data for time series plots was obtained every other month from March 2020 to April of 2021 from the COVID-19 Data Repository by the Center for Systems Science and Engineering (CSSE) at Johns Hopkins University, available on Github at https://github.com/CSSEGISandData/COVID-19 (*17*).

## Acknowledgments

We thank Andrea J. French, Gabriella H. Padovani, Logan Hellinger, Roma Kerby and Shreya Jyotishi for their help with data collection during the summer of 2020 and Arhat Pradhan for technical assistance with creation of supplemental movies. Additionally, we thank Joe Schmid from Symphony RM, for discussion to automate data curation from governor’s websites. Finally, all members of the Lakdawala lab for their critical review of this manuscript.

## Funding statement

This work was funded with internal startup funds from the University of Pittsburgh.

---

**Table S1.**
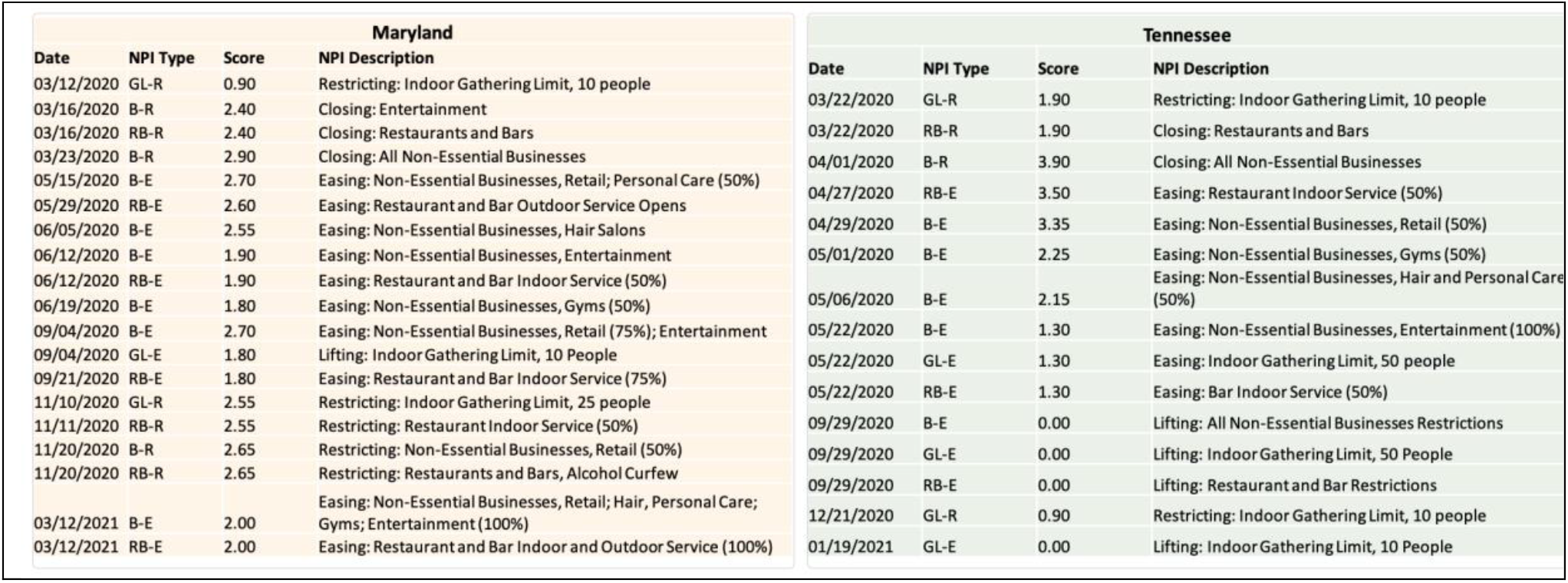
List of NPIs from Maryland and Tennessee by Date

**Table S2:**
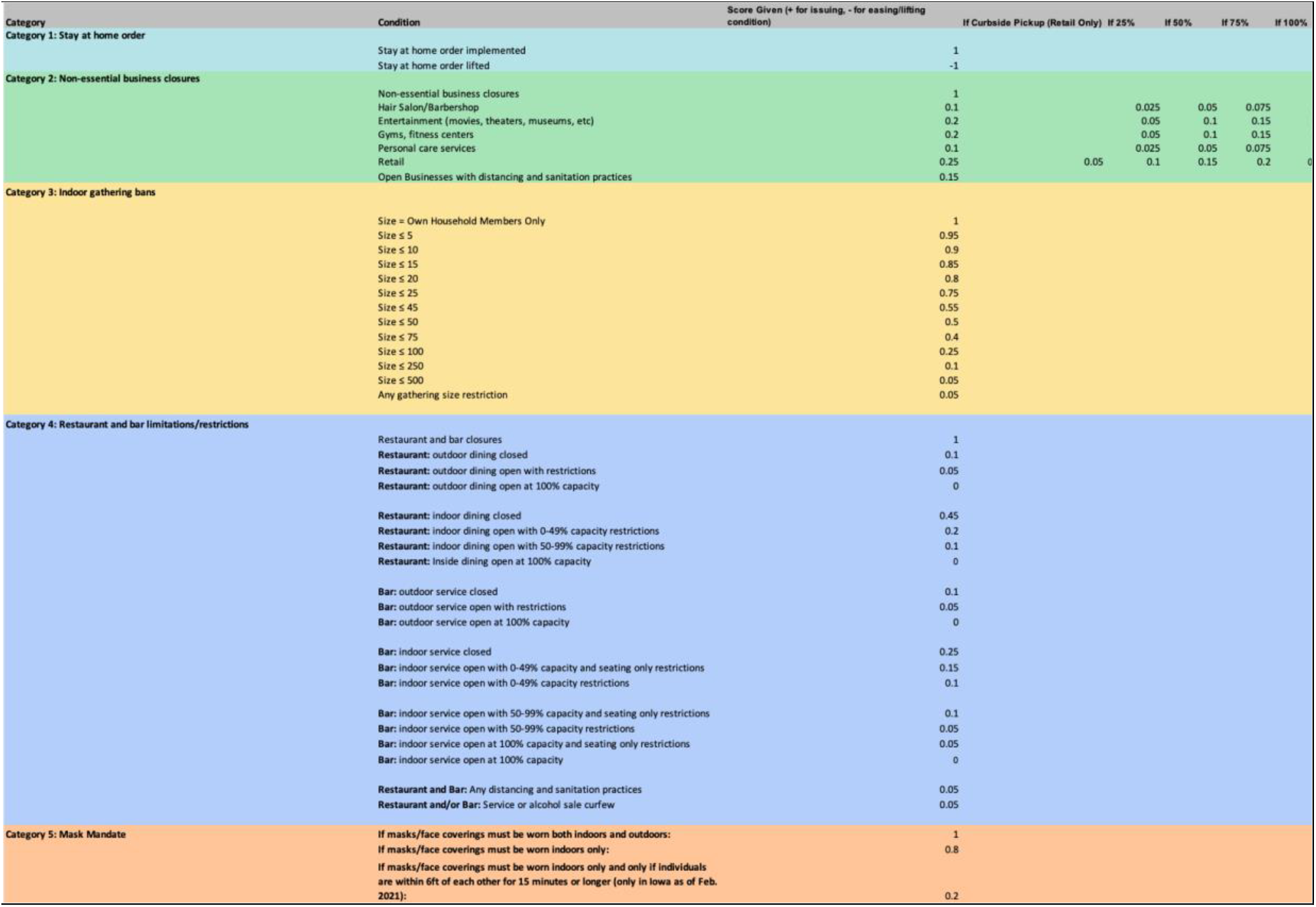
Heuristic NPI Scoring Rubric

**Fig S1:**
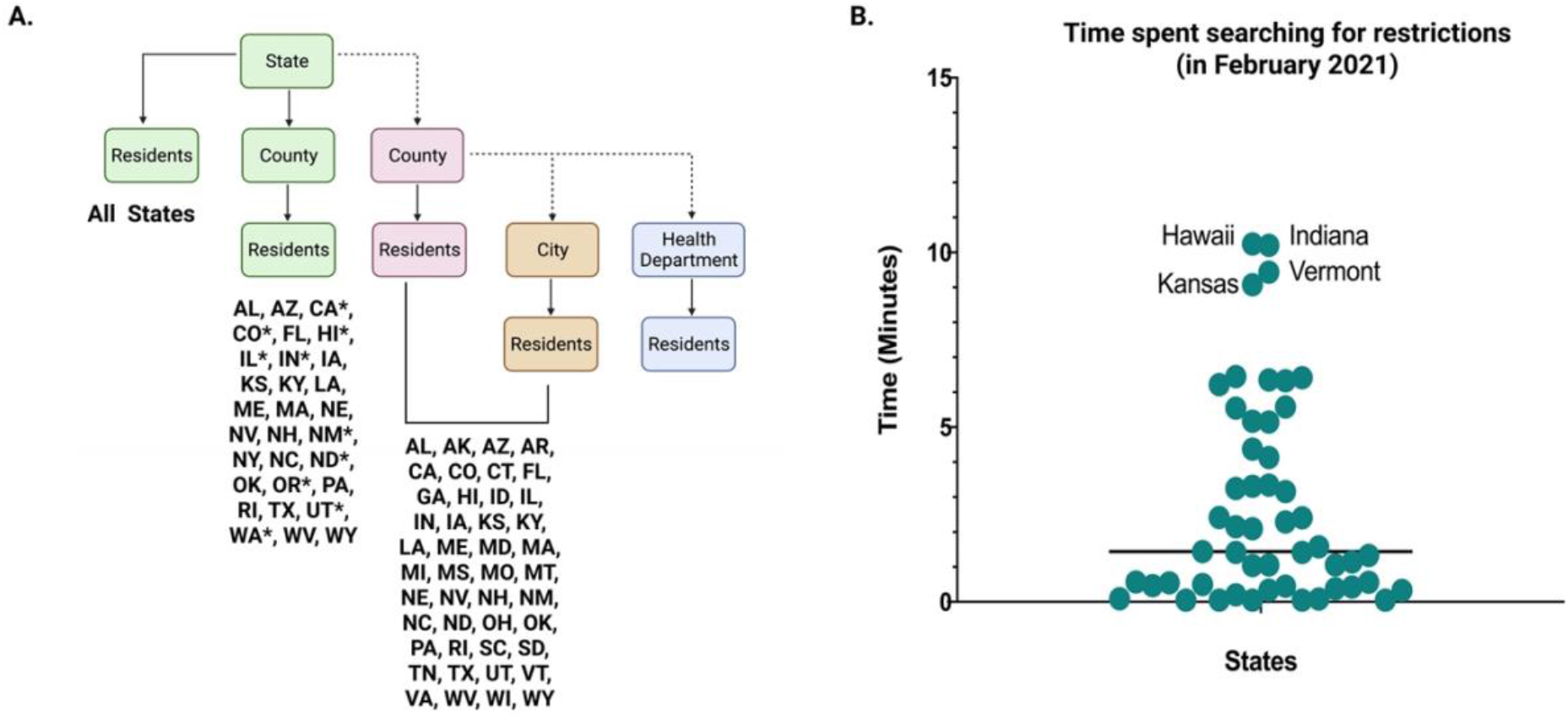
Implementation strategies and accessibility of NPI in each state. (A) State governments issuing NPI on all residents either with entire state orders or county-based orders. Indicated with a star are state governments that adopted a county risk-level metric and regularly changed specific county NPIs depending on the current COVID-19 risk level determined by the state (green). State governments delegating NPI responsibility to counties (red). County governments either delegating NPI responsibilities, or city and health departments deciding to implement NPI on their own (brown and blue respectively). Given the complexity of differentiating between health department and county level implementation, we were unable to capture all the states that implemented NPI through local health departments (B) To quantify accessibility of state COVID-19 NPI information, we timed how long we had to spend searching for NPI information on each state COVID-19 website. Data is from Feb 2021. Some states may have changed the NPI implementation strategy and website for ease.

**Fig S2:**
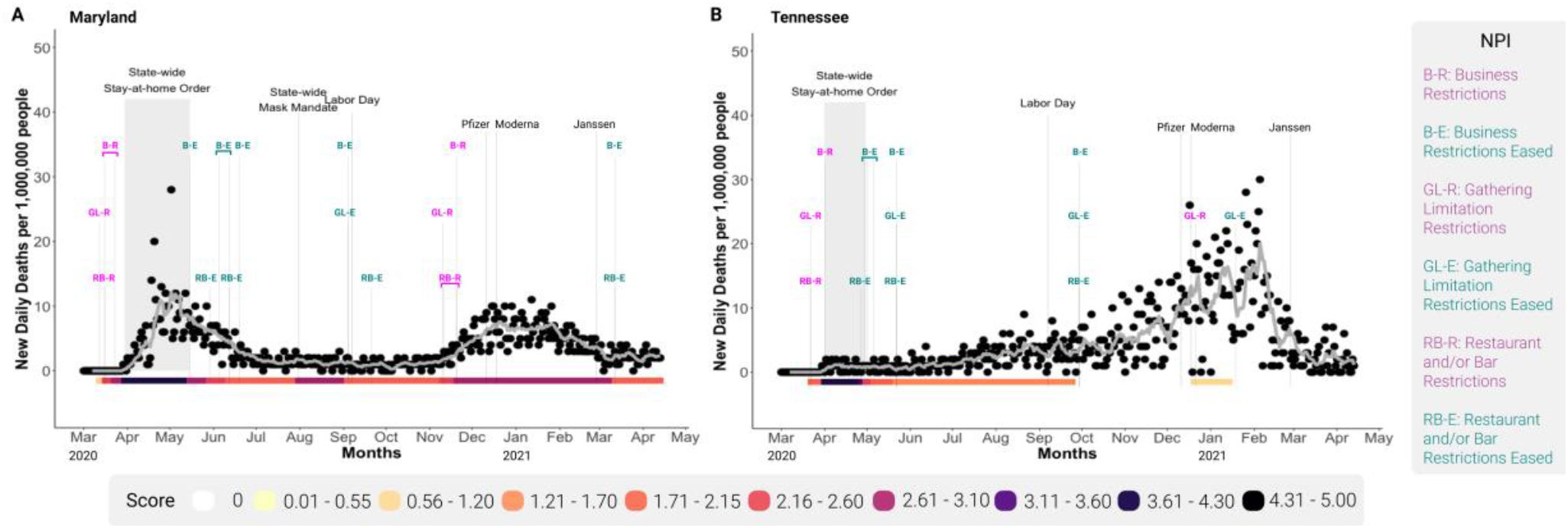
COVID-19 deaths and non-pharmaceutical interventions between states. Time series graphs for Maryland (A) and Tennessee (B) from March 11, 2020 through March 31, 2021. Daily COVID deaths were normalized to state population represented per 1,00,0000 people (black circles) and the 7-day rolling average is indicated by the grey line. State NPI score per day is indicated by the colored bar along the x-axis. The NPIs are labeled with the category abbreviation, and colored as magenta if restricting, or teal if easing. Additionally, the duration of the stay-at-home order is highlighted by the grey band and black lines indicate implementation of statewide mask mandate, labor-day, and each vaccine FDA approval dates.

**Fig S3:**
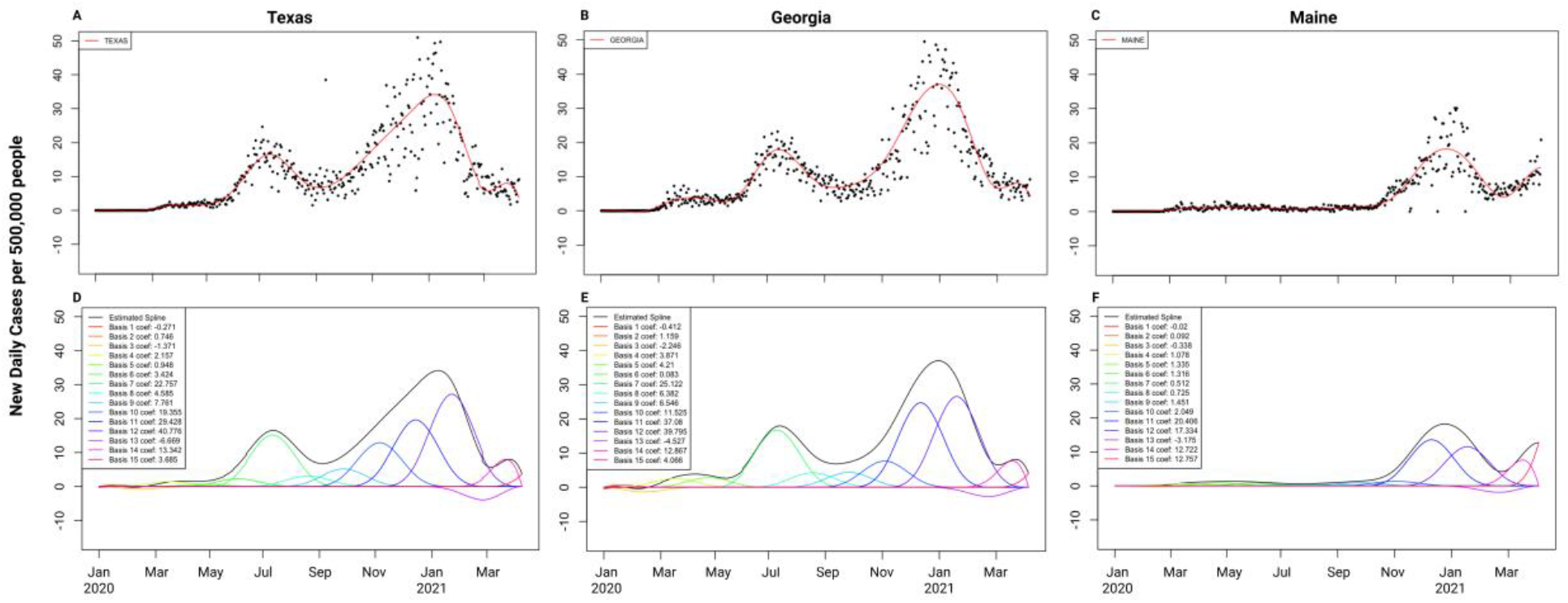
Generation of Basis Spline for Each Sate. Basis spline models were created for the SARS-CoV-2 cases in each state using R package XX, with 15 different knots. (A-C) represents the spline model (red) for three different states (Texas, Georgia, and Maine) on top of each daily case increase. (D-F) Breakdown of the 15 different polynomials that created the spline model and list of the specific coefficient for each spline model.

**Fig S4:**
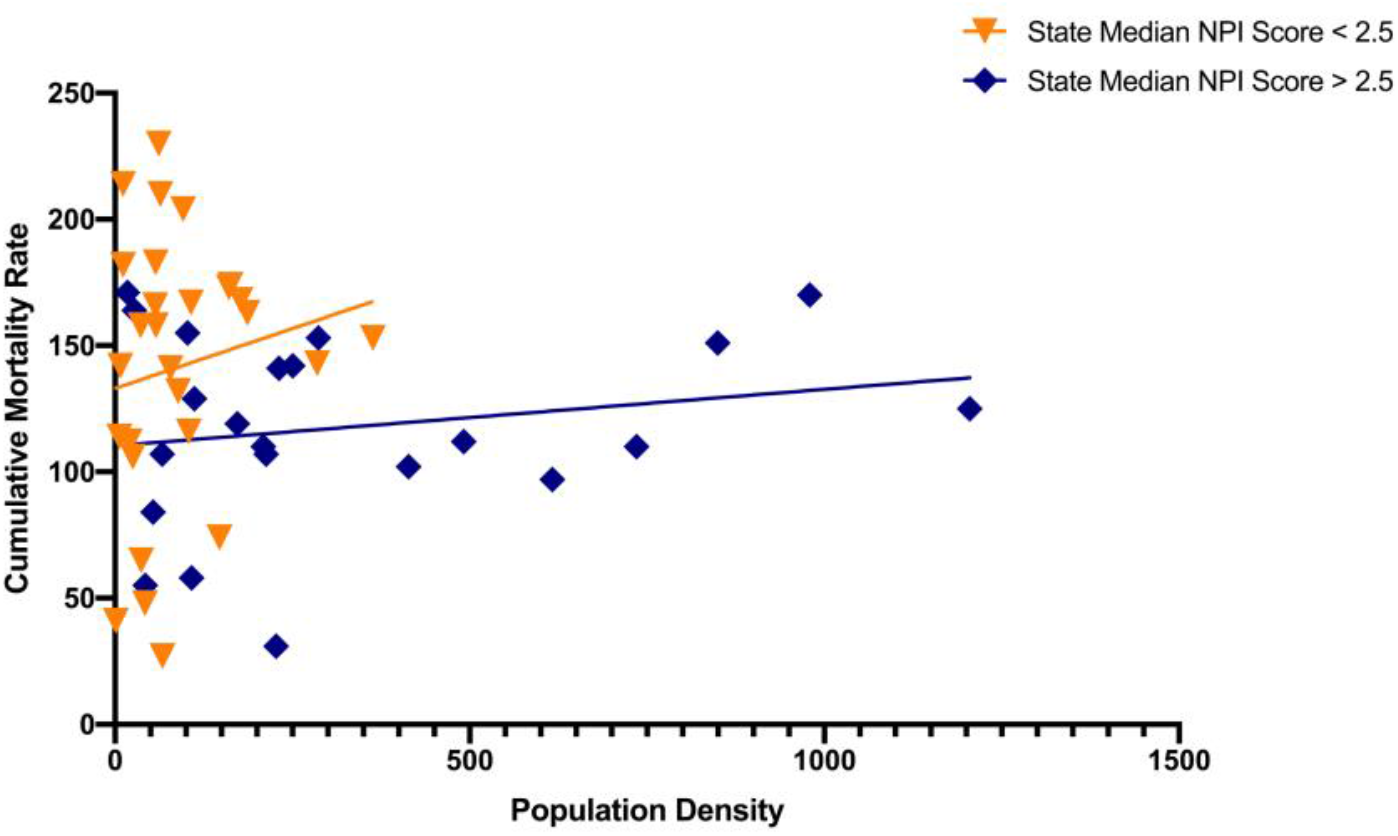
Population density does not correlate with mortality rate. Linear regressions comparing state median score and COVID-19 mortality rate June 1, 2020-April 13, 2021, by state population density.

## Notes

### Competing Interest Statement

The authors have declared no competing interest.

### Author Declarations

This is not a clinical trial and only de-identified SARS-CoV-2 Case data was used for each state.

